# Efficacy and acceptability of an end-user-informed neck resistance training program on outcomes related to sport-related concussion prevention in female football: A quasi-experimental pilot study

**DOI:** 10.64898/2026.11.26.26368814

**Authors:** Branimir Ivanic, Anna Cronström, Nicholas Ryan, Eva Ageberg

**Affiliations:** Department of Health Sciences, Faculty of Medicine, Lund University, Lund, Sweden

## Abstract

**Objectives:** To explore the preliminary efficacy and acceptability of an end-user-informed neck resistance training program on outcomes related to sport-related concussion (SRC) prevention, including linear head acceleration (LHA), rotational head acceleration (RHA), and neck muscle strength.

**Methods:** In this quasi-experimental pilot study, two female football teams were allocated to intervention (n=15) or control (n=15) groups. The intervention group completed a 6-week end-user-informed neck resistance training program, while the control group continued regular training. Peak LHA (g) and RHA (rad/s^2^) were recorded using a motion capture system during standardised head perturbations delivered in multiple directions of cervical motion under anticipated and unanticipated conditions. Neck muscle strength was assessed during maximal voluntary isometric contractions using a handheld dynamometer. Acceptability was assessed using seven Likert-scale items based on the Theoretical Framework of Acceptability. Mixed-design analyses of variance (ANOVA) were used to examine group-by-time interactions, with partial eta squared (η²) reported as the standardised effect-size measure.

**Results:** The intervention group demonstrated significantly greater reductions in both LHA and RHA across anticipatory conditions in the majority of perturbation directions (LHA: p ≤ 0.037, partial η² = 0.17–0.49; RHA: p ≤ 0.016, partial η² = 0.21–0.40), with significantly greater increases in neck muscle strength across all directions (p ≤ 0.003, partial η² = 0.31–0.66). Acceptability ratings were favourable among players and coaches (median range: 3.5–5.0).

**Conclusion:** An end-user-informed neck resistance training program demonstrated preliminary efficacy and favourable acceptability for improving outcomes relevant to SRC prevention in female football players.

**WHAT IS ALREADY KNOWN ON THIS TOPIC:** 1. Female football players experience a high SRC burden but remain under-represented in prevention research.

2. Neck resistance training can improve neck strength, but evidence for reducing head acceleration remains limited

**WHAT THIS STUDY ADDS:** 1. Compared with usual football training, an end-user-informed neck resistance training program was associated with greater reductions in laboratory-measured head acceleration and greater increases in neck strength.

2. The 3–5 minute program was delivered during regular football training using minimal equipment and was rated favourably by players and coaches.

**HOW THIS STUDY MIGHT AFFECT RESEARCH, PRACTICE OR POLICY:** 1. Larger randomised trials with field-based assessments are needed to determine whether laboratory improvements translate to reduced head impact magnitude and SRC risk.

2. Brief, progressive neck training may fit regular football routines, with player and coach feedback guiding adaptations for comfort and seasonal training demands.

## 1. Introduction

Sport-related concussion (SRC) is defined as: “a traumatic brain injury caused by a direct blow to the head, neck or body resulting in an impulsive force being transmitted to the brain that occurs in sports and exercise-related activities” ^1^. Following impact, rapid skull acceleration may cause relative motion between the skull and brain, generating tissue deformation, shear strain, and pressure gradients that may contribute to injury ^2^. As these responses are difficult to measure directly in vivo, head acceleration is commonly used as a surrogate for mechanical loading of the brain ^3^.

Head acceleration comprises two components: linear head acceleration (LHA) and rotational head acceleration (RHA). LHA reflects translational motion and is associated with intracranial pressure changes, whereas RHA reflects angular motion and is associated with brain tissue shear ^2^. Importantly, both have been linked to SRC risk, with concussive impacts in team sports reported to involve approximately 140% greater head acceleration than non-concussive impacts ^4^, highlighting the relevance of reducing head acceleration in sports with a high SRC burden.

Across sporting populations, female athletes often have higher SRC incidence rates than male athletes ^5–7^, with football among the sports with the highest incidence in female athletes ^6–8^. A considerable proportion of SRCs in female football are associated with ball-to-head impacts, particularly during heading ^5^ ^6^. Despite this burden, female athletes remain under-represented in SRC research, accounting for only 37% participants in a recent systematic review and meta-analysis ^9^.

These findings highlight female football as an important setting for SRC prevention and the need to identify modifiable factors that may influence head impact biomechanics and SRC risk.

Alongside improvements in heading technique ^10^, neck muscle strengthening has been proposed to modify head impact biomechanics and reduce SRC risk ^11^. Female football players typically have lower absolute neck muscle strength than male players ^12–14^, and lower strength has been associated with greater head acceleration during heading ^15^ and increased SRC risk ^12^. Greater neck strength may increase stiffness within the head-neck segment, improving head control and attenuating head acceleration following impact ^11^ ^16^. However, a recent systematic review and meta- analysis identified few studies in female football players, with inconsistent effects on head acceleration following neck-strengthening interventions ^17^.

To support implementation in female football, an end-user-informed neck resistance training program was developed with players, coaches, athletic trainers, and physiotherapists ^18^. The primary aim of this study was to explore the program’s preliminary efficacy on outcomes related to SRC prevention (i.e., linear and rotational head acceleration, and neck muscle strength) in female football players. The secondary aim was to evaluate its acceptability.

## 2. Methods

### 2.1. Study design & reporting

This quasi-experimental pilot study was registered at ClinicalTrials.gov (ID: NCT06774976) and reported according to the Transparent Reporting of Evaluations with Nonrandomized Designs (TREND) statement ^19^. The study followed the Declaration of Helsinki ^20^ and was approved by the Swedish Ethical Review Authority (2024-04579-01). All participants provided written informed consent before participation.

### 2.2. Recruitment & allocation

The research team contacted coaches/club representatives of senior female football teams competing in Swedish Divisions 1–4 located in Scania, southern Sweden. Recruitment and allocation were conducted at team level, with one team assigned to intervention and one to control without randomisation. The intervention team was purposively selected to increase the likelihood of successful program implementation during regular football activities.

### 2.3. Participants & sample size

Players were eligible for inclusion if they: 1) were 15 years of age or older; 2) regularly participated in team activities (training and matches); and 3) were expected to be available for team activities during the 6-week intervention period. Players were excluded if they: 1) had any injury to the upper limbs, lower limbs, torso, neck, or head at the time of enrolment; or 2) had a history of neck or head injury within the previous three months.

Given limited prior estimates of head acceleration responses to neck-strengthening interventions, a well-informed sample size calculation was not feasible. Recruitment was therefore limited to two teams to generate preliminary effect-size estimates for future sample size calculations and full-scale trial design.

### 2.4. Intervention

The intervention team was scheduled to complete three sessions per week for 6 weeks, with two sessions omitted due to public holidays, resulting in 16 scheduled sessions in total. The end-user-informed neck resistance training program was developed through three iterative phases, with the detailed development and full exercise content described elsewhere ^18^. Briefly, the program comprised seven exercises organised into three sequential blocks, each combining one rolling static neck activation exercise with one or two dynamic resistance exercises. The program targeted cervical flexion, extension, and left and right rotation, and was performed as a group on the football pitch, requiring 3–5 minutes to complete ^18^.

Assistant coaches delivered the program, with weekly on-site support from the first author (BI) and regular communication between sessions. Progression was incorporated by increasing the duration of the rolling static exercises at the team level and individualised resistance for the dynamic exercises using a head harness with adjustable resistance tubing (*Alpha Plus Super Bundle, Iron Neck, USA*). Resistance was increased when participants completed the prescribed 8–12 repetitions with correct form and without discomfort.

Before commencement of the 6-week intervention period, the research team delivered an educational presentation to the intervention group covering SRC, the biomechanical rationale for the neck resistance training, and the potential role of increased cervical strength in reducing head acceleration and SRC risk. The presentation also included instructions on correct exercise technique and safety considerations to reduce the risk of discomfort or injury during training.

### 2.5. Control

At the player and coach information meeting, the control team received general information on SRC, its incidence in female football, female athletes’ under-representation in SRC research, and the relevance of head acceleration to SRC risk. The study was presented as an evaluation of head acceleration changes following six weeks of regular preseason football training. Consequently, the control team was unaware that an intervention was being evaluated in another team. The control team continued their regular preseason training, with the first author maintaining weekly contact with the head coach to monitor training continuity.

### 2.6. Assessment

#### 2.6.1. Settings

Laboratory assessments took place between January and May 2026. Both groups completed baseline and follow-up assessments of head acceleration and neck muscle strength in the same motion analysis room using the same equipment, testing setup, and standardized protocol. All sessions were conducted by the first author together with one of three trained project assistants, who rotated between sessions.

In the intervention group, acceptability questionnaires were completed at the follow-up laboratory visit. Assistant coached recorded adherence and exercise progression during training throughout the 6-week intervention. In addition, players were instructed to report any intervention-related adverse events, including neck muscle soreness, headache, or other head or neck symptoms, to the coaches.

#### 2.6.2. Participant characteristics

Age was recorded for each participant. Standing height was assessed using a wall-mounted stadiometer, while body mass was measured using a digital weighing scale. Neck circumference was measured at the level of the thyroid cartilage using a flexible tape. Head mass was measured to estimate the head’s centre of mass. For this procedure, passive head weight was assessed using a handheld dynamometer (*Series 3 Digital Force Gauge, Mark-10 Corporation, USA*), with participants supine, the head beyond the table edge and neck muscles relaxed.

#### 2.6.3. Outcomes

##### Head acceleration testing

Prior to head acceleration testing, wireless surface electromyography (sEMG) sensors were placed bilaterally over the sternocleidomastoid muscles and neck extensors below the occipital hairline, using established anatomical landmarks ^21^ ^22^.

Participants then completed a preparatory verification procedure while seated, with instructions to relax their neck muscles or brace as if anticipating ball or player contact. sEMG signals were visually monitored in real time to confirm greater amplitude during activation than relaxation.

Head acceleration was assessed using a custom-built laboratory apparatus developed to apply controlled, brief mechanical loads to the head and neck. A full technical description of the apparatus and testing procedure has been reported elsewhere ^23^. In brief, participants sat with the upper torso restrained and wore a head harness connected via a taut steel wire to a pulley system. A 1-kg weight was released from a standardised drop height adjusted to participant body mass, transmitting a brief, controlled tensile force to the head.

Trials were conducted in flexion, extension, right lateral flexion, and left rotation, in randomised order. For each direction, participants completed one familiarisation trial followed by three recorded trials under both anticipated and unanticipated conditions. During anticipated trials, participants were instructed to maximally pre-activate the neck muscles before load release; during unanticipated trials, they wore an eye mask and were instructed to relax. sEMG was monitored in real time to verify the intended activation condition. This protocol has demonstrated moderate-to-excellent test-retest reliability (ICC 0.72 – 0.90) across movement directions and anticipatory conditions ^23^.

Head kinematics were recorded at 100 Hz using a three-dimensional motion capture system (*Qualisys AB, Gothenburg, Sweden*). Reflective markers tracked head motion, while a marker on the on the falling weight identified load onset. Data were processed in MATLAB (*MathWorks, Natick, USA*) to extract peak LHA (*g*), and RHA (*rad/s*^2^) for each trial.

##### Neck muscle strength

The neck muscle strength testing protocol was informed by published guidance on neck strength assessment for sports medicine clinicians ^24^. Before maximal voluntary isometric contraction (MVIC) testing, participants completed a 1-minute warm-up of dynamic movements through the full range of motion in all cervical directions.

The head harness was connected to a handheld dynamometer secured to wall-mounted metal bars. Participants sat upright on a bench with the torso neutral and hands resting on thighs. Straps stabilised the upper torso to minimise trunk contribution, while the first author monitored head, neck, and trunk alignment.

Participants were instructed to avoid ballistic movements, gradually build force over 3 seconds and maintain maximal effort for 2 seconds. Following one familiarisation trial per direction, participants completed two MVIC trials in flexion, extension, left lateral flexion, and right rotation, in randomised order. The first author provided verbal encouragement, with 30-second rests between trials. An additional trial was performed if the first two differed by >10%, and the highest value per direction was retained for analysis.

MVIC force (N) was converted to moments (N·m) by multiplying each force by the estimated neck moment arm. Calculations assumed fixed axes of rotation at C7 for flexion, extension, and lateral flexion ^25^ ^26^, and at the midpoint between the ears for rotation ^22^.

### Acceptability questionnaire

Program acceptability was assessed among players and coaches after the 6-week intervention using a questionnaire based on the Theoretical Framework of Acceptability ^27^. It included seven statements rated on a 5-point Likert scale (1 = strongly disagree to 5 = strongly agree) and two open-ended questions (*Supplemental material 1*).

### 2.7. Statistical analyses

Analyses were performed using Stata 19.5 (*StataCorp LLC, College Station, USA*). Trials affected by visually identified motion artefacts were excluded, and distributions were visually inspected for normality. Participant characteristics, as well as baseline, follow-up, and change-from-baseline values for LHA, RHA, and neck strength outcomes were summarised as means and standard deviations. Change scores were calculated as follow-up minus baseline, while composite scores as the mean across the four directions for each outcome.

Group-by-time interactions were examined using mixed-design analyses of variance (ANOVA), with group (intervention vs. control) as the between-subject factor and time (baseline vs. follow-up) as the within-subject factor. Analyses were performed separately for LHA and RHA in each perturbation direction (flexion, extension, right lateral flexion, left rotation, and composite score) and condition (anticipated and unanticipated); and each neck strength direction (flexion, extension, left lateral flexion, right rotation, and composite score).

Between-group differences in change were reported with 95% confidence intervals (CIs) and p-values. Partial eta squared (η²) with 95% CIs was calculated for group-by-time interactions and interpreted as small (0.01 – 0.05), medium (0.06 – 0.13), or large (≥ 0.14) ^28^. Statistical significance was set at p < 0.05. No correction for multiple comparisons was applied given the exploratory pilot design; findings were interpreted cautiously alongside effect sizes.

Acceptability questionnaire data were summarised descriptively. Negatively worded items were reverse-scored before analysis, with higher scores indicating greater acceptability. Likert responses were summarised as median and interquartile range for players and median and range for coaches, given the small coach sample. Open-ended were summarised narratively to describe barriers and suggestions for continued use.

### 2.8. Patient & public involvement

A subgroup of players and assistant coaches from the intervention team contributed directly to refinement of the neck resistance training program during the preceding developmental study ^18^. Players and coaches also informed the implementation strategy, including decisions on timing within the football season, session frequency and the organisation of training groups. Their involvement in decision-making helped align the program delivery with the team’s routines and practical constraints, supporting implementation during regular football activities.

### 2.9. Equality, diversity & inclusion

The author team comprised two male and two female researchers and represents a combination of junior (2 authors) and senior (2 authors) researchers from Lund University, Sweden. The study focused on female football players, who experience a high SRC burden yet remain under-represented in SRC prevention research. This focus was intended to address sex-related gaps in the evidence and support more equitable SRC prevention research and practice.

## 3. Results

### 3.1. Participant flow

Of 9 female football teams contacted, 2 agreed to participate and 7 did not respond. Of 40 players invited, 30 (75%) consented and completed baseline testing, while 10 (5 per group) did not volunteer. Three players were lost to follow-up (1 intervention, 2 control) for reasons unrelated to the intervention, leaving 27 (90%) in the final analysis (Figure 1).

**Figure 1.**
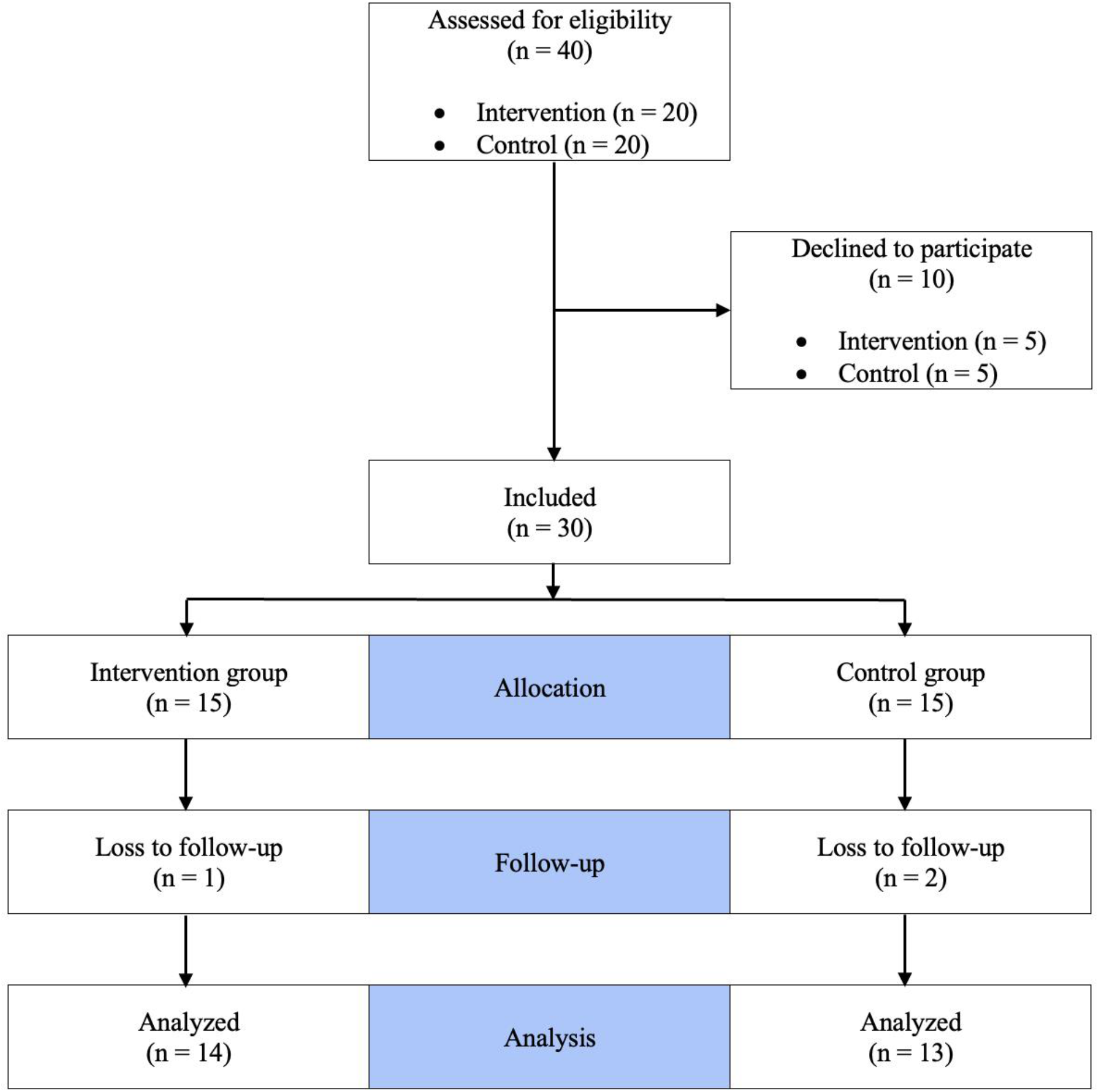
Flow of participants from eligibility assessment to final analysis.

### 3.2. Baseline characteristics

The intervention and control teams competed in Swedish Divisions 3 and 4, respectively. Intervention players were younger than control players (16.9 ± 0.6 vs. 22.0 ± 6.4 years), while other baseline characteristics were comparable between groups (Table 1).

**Table 1.** Characteristics of included participants.

|  | **Intervention**<br>Mean (SD)<br>n = 14 | **Control**<br>Mean (SD)<br>n = 13 | **All participants**<br>Mean (SD)<br>n = 27 |
|---|---|---|---|
| **Age** (yrs) | 16.9 (0.6) | 22.0 (6.4) | 19.3 (5.1) |
| **Height** (cm) | 166.6 (4.6) | 167.9 (6.9) | 167.3 (5.8) |
| **Body mass** (kg) | 65.2 (10.1) | 67.9 (10.0) | 66.5 (9.9) |
| **Head mass** (kg) | 3.8 (0.4) | 3.8 (0.5) | 3.8 (0.4) |
| **Neck circumference** (cm) | 32.3 (1.9) | 32.4 (1.9) | 32.3 (1.9) |

***SD.*** Standard Deviation.

### 3.3. Adherence, progression, & adverse events

Mean adherence was 11.2 sessions per player (70%; range 8–14 sessions, 50% – 88%). Weekly adherence averaged 1.9 sessions per player, with weekly means ranging from 1.3 to 2.3 sessions. Rolling static exercises progressed at team level from 30 to 60 seconds by week 6. All players started with 5-lb resistance tubing in week 1, progressing to 10-lb during weeks 2–3 and 25-lb tubing during weeks 3–4. By week 4, all were using 25-lb tubing, maintained through week 6. No intervention-related adverse events were reported.

### 3.4. Head acceleration

#### 3.4.1. Linear head acceleration

During anticipated trials, the intervention group demonstrated significantly greater reductions in LHA for flexion, extension, rotation, and composite score, with between-group differences in change ranging from -0.06 to -0.03 g (p values ≤0.018; partial η² = 0.21–0.44). No significant between-group difference was observed for lateral flexion (p=0.256; Table 2).

**Table 2.** Comparison of LHA from baseline to follow-up between the intervention (n = 14) and control (n = 13) groups.

| **Direction** | **Condition** | **Intervention**<br>Mean (SD) |  |  | **Control**<br>Mean (SD) |  |  | **Mean difference in change**<br>(95% CI) | **p value** | **η²**<br>(95% CI) |
|---|---|---|---|---|---|---|---|---|---|---|
|  |  | Baseline | Follow-up | Change | Baseline | Follow-up | Change |  |  |  |
| **Flexion** | Anticipated | 0.23<br>(0.03) | 0.19<br>(0.02) | -0.04<br>(0.03) | 0.24<br>(0.03) | 0.23<br>(0.03) | -0.01<br>(0.03) | -0.03<br>(-0.06 to -0.01) | **0.009** | 0.25<br>(0.02 to 0.48) |
|  | Unanticipated | 0.25<br>(0.03) | 0.21<br>(0.05) | -0.04<br>(0.03) | 0.24<br>(0.03) | 0.25<br>(0.04) | 0.01<br>(0.04) | -0.05<br>(-0.08 to -0.02) | **0.002** | 0.33<br>(0.06 to 0.54) |
| **Extension** | Anticipated | 0.24<br>(0.03) | 0.20<br>(0.02) | -0.04<br>(0.03) | 0.23<br>(0.03) | 0.25<br>(0.04) | 0.02<br>(0.05) | -0.06<br>(-0.09 to -0.02) | **0.002** | 0.32<br>(0.05 to 0.54) |
|  | Unanticipated | 0.27<br>(0.05) | 0.21<br>(0.03) | -0.06<br>(0.04) | 0.28<br>(0.05) | 0.28<br>(0.05) | 0.00<br>(0.05) | -0.05<br>(-0.09 to -0.02) | **0.005** | 0.29<br>(0.03 to 0.51) |
| **Lateral flexion** | Anticipated | 0.22<br>(0.03) | 0.19<br>(0.03) | -0.03<br>(0.03) | 0.24<br>(0.02) | 0.22<br>(0.02) | -0.01<br>(0.04) | -0.02<br>(-0.04 to 0.01) | 0.256 | 0.05<br>(0.00 to 0.27) |
|  | Unanticipated | 0.25<br>(0.03) | 0.23<br>(0.03) | -0.03<br>(0.03) | 0.26<br>(0.03) | 0.26<br>(0.03) | 0.00<br>(0.04) | -0.03<br>(-0.06 to -0.02) | **0.037** | 0.17<br>(0.01 to 0.41) |
| **Rotation** | Anticipated | 0.15<br>(0.03) | 0.13<br>(0.02) | -0.03<br>(0.03) | 0.16<br>(0.02) | 0.16<br>(0.02) | 0.00<br>(0.02) | -0.03<br>(-0.05 to -0.01) | **0.018** | 0.21<br>(0.01 to 0.45) |
|  | Unanticipated | 0.18<br>(0.03) | 0.15<br>(0.03) | -0.04<br>(0.04) | 0.18<br>(0.04) | 0.19<br>(0.05) | 0.01<br>(0.04) | -0.05<br>(-0.08 to -0.01) | **0.007** | 0.26<br>(0.02 to 0.49) |
| **Composite** | Anticipated | 0.21<br>(0.02) | 0.18<br>(0.01) | -0.03<br>(0.02) | 0.22<br>(0.01) | 0.22<br>(0.02) | 0.00<br>(0.02) | -0.03<br>(-0.05 to -0.02) | **< 0.001** | 0.44<br>(0.14 to 0.62) |
|  | Unanticipated | 0.24<br>(0.02) | 0.20<br>(0.02) | -0.04<br>(0.02) | 0.24<br>(0.02) | 0.25<br>(0.02) | 0.00<br>(0.03) | -0.05<br>(-0.07 to -0.03) | **< 0.001** | 0.49<br>(0.19 to 0.66) |

LHA values presented in *g*. Bolded values indicate statistically significant differences (p < 0.05). ***SD.*** Standard Deviation; ***η².*** Partial eta squared.

During unanticipated trials, significantly greater reductions occurred in the intervention group across all tested directions and composite score, with between-group differences in change ranging from -0.05 to -0.03 g (p values ≤0.037; partial η² = 0.17–0.49; Table 2).

#### 3.4.2. Rotational head acceleration

During anticipated trials, the intervention group demonstrated significantly greater reductions in RHA for flexion, extension, rotation, and composite score, with between-group differences in change ranging from -6.32 to -3.66 rad/s^2^ (p values ≤0.016; partial η² = 0.21–0.25. No significant between-group difference was observed for lateral flexion (p =0.075; Table 3).

**Table 3.** Comparison of RHA from baseline to follow-up between the intervention (n = 14) and control (n = 13) groups.

| **Direction** | **Condition** | **Intervention**<br>Mean (SD) |  |  | **Control**<br>Mean (SD) |  |  | **Mean difference in change**<br>(95% CI) | **p value** | **η²**<br>(95% CI) |
|---|---|---|---|---|---|---|---|---|---|---|
|  |  | Baseline | Follow-up | Change | Baseline | Follow-up | Change |  |  |  |
| **Flexion** | Anticipated | 22.99<br>(3.47) | 19.58<br>(2.30) | -3.44<br>(3.63) | 23.19<br>(3.39) | 23.52<br>(4.05) | -0.33<br>(3.22) | -3.76<br>(-6.49 to -1.03) | **0.008** | 0.24<br>(0.02 to 0.47) |
|  | Unanticipated | 18.46<br>(3.27) | 14.78<br>(2.21) | -3.69<br>(3.11) | 20.12<br>(3.71) | 20.69<br>(4.14) | 0.57<br>(4.89) | -4.25<br>(-7.48 to -1.03) | **0.012** | 0.23<br>(0.01 to 0.46) |
| **Extension** | Anticipated | 28.42<br>(3.86) | 24.38<br>(5.49) | -4.03<br>(4.96) | 24.62<br>(5.25) | 26.91<br>(4.79) | 2.29<br>(7.28) | -6.32<br>(-11.36 to -1.28) | **0.016** | 0.22<br>(0.01 to 0.45) |
|  | Unanticipated | 32.06<br>(8.37) | 26.17<br>(4.78) | -5.89<br>(7.73) | 31.09<br>(4.99) | 34.41<br>(8.04) | 3.31<br>(7.07) | -9.20<br>(-15.20 to -3.21) | **0.004** | 0.29<br>(0.04 to 0.52) |
| **Lateral flexion** | Anticipated | 21.12<br>(5.20) | 20.00<br>(4.27) | -1.12<br>(4.13) | 18.59<br>(3.48) | 20.43<br>(4.72) | 1.84<br>(3.98) | -2.96<br>(-6.24 to 0.32) | 0.075 | 0.13<br>(0.00 to 0.37) |
|  | Unanticipated | 23.39<br>(4.80) | 21.06<br>(5.53) | -2.32<br>(5.09) | 22.37<br>(4.73) | 25.33<br>(5.93) | 2.96<br>(2.14) | -5.28<br>(-8.44 to -2.13) | **0.002** | 0.33<br>(0.06 to 0.55) |
| **Rotation** | Anticipated | 40.67<br>(4.61) | 37.35<br>(5.09) | -3.32<br>(4.67) | 39.23<br>(3.91) | 40.52<br>(5.09) | 1.29<br>(4.49) | -4.61<br>(-8.24 to -0.97) | **0.015** | 0.21<br>(0.01 to 0.45) |
|  | Unanticipated | 43.62<br>(2.55) | 38.14<br>(5.41) | -5.48<br>(5.38) | 42.85<br>(4.58) | 43.67<br>(6.19) | 1.61<br>(7.11) | -7.09<br>(-12.20 to -1.98) | **0.009** | 0.25<br>(0.02 to 0.49) |
| **Composite** | Anticipated | 28.66<br>(3.38) | 25.74<br>(3.00) | -2.91<br>(3.37) | 26.99<br>(2.42) | 27.74<br>(0.74) | 0.74<br>(3.21) | -3.66<br>(-6.27 to -1.05) | **0.008** | 0.25<br>(0.02 to 0.48) |
|  | Unanticipated | 28.78<br>(4.70) | 24.78<br>(3.54) | -4.00<br>(3.99) | 29.13<br>(3.76) | 31.14<br>(4.16) | 2.00<br>(3.54) | -6.01<br>(-9.01 to -3.01) | **< 0.001** | 0.40<br>(0.11 to 0.60) |

RHA values presented in *rad/s<sup>2</sup>*. Bolded values indicate statistically significant differences (*p* < 0.05). ***SD.*** Standard Deviation; ***η².*** Partial eta squared.

During unanticipated trials, significantly greater reductions occurred in the intervention group across all tested directions and composite score, with between-group differences in change ranging from -9.20 to -4.25 rad/s^2^ (p values ≤0.012; partial η² = 0.23–0.40; Table 3).

### 3.5. Neck muscle strength

The intervention group demonstrated significantly greater improvements in all tested neck strength directions and in the composite score, with between-group differences in change ranging from 3.24 to 4.01 N·m (p values ≤0.003; partial η² = 0.31–0.66; Table 4).

**Table 4.** Comparison of neck muscle strength from baseline to follow-up between the intervention (n = 14) and control (n = 13) groups.

| **Direction** | **Intervention**<br>Mean (SD) |  |  | **Control**<br>Mean (SD) |  |  | **Mean difference in change**<br>(95% CI) | **p value** | **η²**<br>(95% CI) |
|---|---|---|---|---|---|---|---|---|---|
|  | Baseline | Follow-up | Change | Baseline | Follow-up | Change |  |  |  |
| **Flexion** | 13.87<br>(3.89) | 17.80<br>(3.95) | 3.93<br>(2.03) | 12.1<br>(3.79) | 12.02<br>(3.64) | -0.08<br>(0.49) | 4.01<br>(2.82 to 5.20) | **< 0.001** | 0.66<br>(0.39 to 0.78) |
| **Extension** | 17.84<br>(3.30) | 21.93<br>(3.40) | 4.09<br>(2.61) | 18.30<br>(4.80) | 18.41<br>(3.86) | 0.12<br>(0.36) | 3.98<br>(2.47 to 5.48) | **< 0.001** | 0.54<br>(0.24 to 0.70) |
| **Lateral flexion** | 15.98<br>(3.14) | 19.93<br>(4.46) | 3.95<br>(4.01) | 13.48<br>(3.72) | 13.67<br>(3.73) | 0.20<br>(0.37) | 3.75<br>(1.45 to 6.05) | **0.003** | 0.31<br>(0.05 to 0.53) |
| **Rotation** | 9.53<br>(1.91) | 12.92<br>(2.03) | 3.39<br>(2.28) | 8.35<br>(1.61) | 8.50<br>(1.56) | 0.15<br>(0.46) | 3.24<br>(1.91 to 4.57) | **< 0.001** | 0.50<br>(0.20 to 0.67) |
| **Composite** | 14.30<br>(2.58) | 18.14<br>(2.51) | 3.84<br>(1.96) | 13.06<br>(2.82) | 13.15<br>(2.78) | 0.10<br>(0.22) | 3.75<br>(2.62 to 4.87) | **< 0.001** | 0.65<br>(0.38 to 0.77) |

Neck muscle strength values presented in *N·m*. Bolded values indicate statistically significant differences (p < 0.05). ***SD.*** Standard Deviation; ***η².*** Partial eta squared.

### 3.6. Acceptability of the program

Acceptability ratings were favourable after the 6-week intervention with median item scores of 4–5 for players and 3.5–4.5 for coaches. Players generally reported liking the program, confidence in performing it and understanding its potential relevance for SRC prevention, alongside limited discomfort, manageable effort, and little interference with other priorities. Coaches rated supervising and implementing the program positively (Table 5).

**Table 5.** Player and coach acceptability ratings of the end-user-informed neck resistance training program following the 6-week intervention.

|  | **Player statements (n = 14)** | **Median (IQR)** |
|---|---|---|
| **1.** | *I liked the exercise program.* | 4 (4–4) |
| **2.** | *I felt uncomfortable while engaging in the exercise program.* | 5 (4–5) |
| **3.** | *This exercise program requires too much effort to perform.* | 4 (4–5) |
| **4.** | *I could benefit from continuing to use this exercise program multiple times per week.* | 4 (4–4) |
| **5.** | *It is clear to me how this exercise program may affect outcomes related to prevention of sport–related concussion.* | 4 (4–5) |
| **6.** | *I felt confident performing the exercise program.* | 5 (5–5) |
| **7.** | *Implementing this exercise program interfered with my other priorities.* | 4 (4–5) |
|  | **Coach statements (n = 2)** | **Median (range)** |
| **1.** | *I liked the exercise program.* | 4.5 (4–5) |
| **2.** | *I felt uncomfortable while supervising the exercise program.* | 4.5 (4–5) |
| **3.** | *This exercise program requires too much effort to organize.* | 4 (4–4) |
| **4.** | *Our team could benefit from continuing to use this exercise program multiple times per week.* | 3.5 (3–4) |
| **5.** | *It is clear to me how this exercise program may affect outcomes related to prevention of sport–related concussion.* | 4 (4–4) |
| **6.** | *I felt confident supervising the exercise program.* | 4.5 (4–5) |
| **7.** | *Implementing this exercise program interfered with my other priorities.* | 4 (4–4) |

Negatively worded items were reverse-scored so that higher scores indicate greater acceptability for all items. ***IQR.*** Interquartile range.

In open-ended responses, several players identified rolling static exercises as the main challenge for continued use. They reported greater discomfort on natural than artificial grass and suggested placing a thin exercise mat under the upper back. Coaches identified maintaining the program at every training session throughout the competitive season as the main challenge. They suggested implementation at every preseason training session, followed by a maintenance once per week during the season.

## 4. Discussion

This quasi-experimental pilot study provides preliminary support for the program’s efficacy in reducing head acceleration and improving neck muscle strength. Reductions in LHA and RHA were significantly greater in the intervention group than in control group under anticipated and unanticipated conditions, except for anticipated lateral flexion. Neck muscle strength improvements were also significantly greater in the intervention group across all directions and the composite strength score. Estimated effect sizes were large for all statistically significant comparisons. Finally, favourable player and coach ratings suggested that the program was perceived as relevant, understandable, and feasible for team-based implementation.

### 4.1. Head acceleration & neck muscle strength

The intervention group demonstrated greater reductions in head acceleration across most comparisons. These reductions were evident for both LHA and RHA, indicating effects across both components of head motion. This is particularly relevant because sport-related head impacts, and the resulting mechanical loading of the brain, typically involve combined linear and rotational components ^29^. The consistency across both outcomes therefore provides encouraging preliminary support for the potential of the end-user-informed neck resistance training program to influence the head’s overall biomechanical response to external perturbations.

Reductions in head acceleration were also observed under both anticipated and unanticipated conditions. This is notable because many sport-related impacts occur when athletes have limited awareness and time to prepare for contact ^30^. Unanticipated collisions generally result in greater head acceleration than anticipated impacts ^31^. Consistent with previous research ^23^, head accelerations in the present study were higher during unanticipated trials, likely reflecting less opportunity to activate the neck musculature and stabilise the head before perturbation. However, these controlled laboratory perturbations may not fully replicate impacts during football play, supporting future evaluation under more ecologically valid field-based conditions.

Although neck resistance training can improve cervical muscle strength, evidence regarding its effects on head acceleration remains limited ^17^. In the present study, a short, progressive, multidirectional program was accompanied by significant improvements in neck strength and reductions in laboratory-measured head acceleration, adding preliminary evidence in this area. The concurrent strength gains provide a plausible physiological basis for the acceleration reductions, as greater cervical force-generating capacity may improve head stabilisation and resistance to external perturbation ^11^. This may be especially relevant during anticipated trials, when participants could brace through cervical muscle activation. At the same time, reductions in head acceleration during unanticipated trials suggest that the program’s potential benefits may not depend exclusively on deliberate pre-impact bracing. One possible explanation is that greater cervical force-generating capacity may attenuate the head’s kinematic response independently of anticipatory muscle activation, consistent with previous associations between greater neck muscle strength and reduced head motion under both relaxed and activated conditions ^32^.

Taken together, the parallel strength gains and head acceleration reductions provide a coherent and encouraging pattern. Larger randomised studies are required to confirm these findings and determine whether they translate to reduced head impact magnitude during football participation.

### 4.2. Acceptability of the program

Acceptability of the neck resistance training program was generally favourable among both players and coaches. This finding supports the value of involving end-users early in intervention planning and development, as adoption and sustained implementation of sports injury prevention programs depend not only on efficacy, but also on how intended users perceive the program’s relevance and fit within their everyday sporting context ^33^.

Players reported different experiences of rolling exercises on artificial and natural grass, highlighting a practical issue that may influence comfort and usability. Players also suggested practical solutions, reinforcing the value of involving athletes directly in intervention development and evaluation. These observations support involving multiple end-user groups, as players and coaches may identify different factors influencing the implementation and maintenance of injury prevention program ^34^.

Coaches anticipated that sustaining the program throughout the competitive season would be the main implementation challenge. Their suggestion to perform the program more frequently during preseason, followed by a lower-frequency maintenance phase during the competitive season, aligns with previous football injury prevention approaches ^35^ ^36^. Such a phased approach may support longer- term implementation by maintaining a training stimulus while reducing the perceived burden during periods with greater football-specific demands.

### 4.3. Clinical implications

This brief program may offer clinicians and coaches a practical option for integrating progressive neck strengthening into regular female football training. Minimal equipment requirements and favourable player and coach ratings support its potential fit withing team routines. However, the observed laboratory improvements do not establish reduced SRC risk. Larger randomised trials incorporating field-based head impact assessments and SRC surveillance are needed to determine its role in SRC prevention.

### 4.4. Strengths & limitations

A key strength of this study was that the use of an end-user-informed neck resistance training program within a real-world team environment. The program was also grounded in resistance-training principles, incorporating individual progression in external resistance and a multidirectional combination of static and dynamic exercises. Another strength was the use of objective laboratory-based testing of head acceleration and neck muscle strength using standardised and reliable procedures. Compared with on-field testing, this approach provided greater control over testing conditions and reduced uncontrolled variation related to impact exposure and situational factors, allowing a more focused evaluation of the outcomes of interest.

Several limitations should be acknowledged. First, the quasi-experimental design, with one team per group and no randomisation, limits causal inference and introduces potential selection bias. The pilot nature and small sample size also limit the generalisability. This design was chosen to evaluate preliminary efficacy while increasing the likelihood of successful implementation. Second, laboratory perturbations were considerably smaller than many real-world football impacts. Mean LHA and RHA in the present study were 0.25 g and 28.39 rad/s^2^, respectively, compared with 27.80 g and 3280 rad/s^2^ for real-world non-concussive football impacts reported in a recent systematic review ^4^. Although this limits ecological validity, the protocol was designed to prioritise participant safety while allowing standardised assessment of head acceleration responses. Finally, SRC incidence was not assessed, so the findings represent changes in surrogate biomechanical and neuromuscular outcomes rather than direct evidence of SRC prevention.

## 5. Conclusion

The findings provide preliminary support for the efficacy of an end-user-informed neck resistance training program in improving outcomes relevant to SRC prevention in female football players, including reduced laboratory-measured head acceleration and increased neck muscle strength. Favourable acceptability ratings from players and coaches support its potential for integration into a team-based football environment. Larger randomised controlled trials are needed to further evaluate these effects and determine whether they translate to reduced head impact magnitude and SRC risk during football participation.

## Other information

### Contributorship statement

BI was involved in the conceptualization of the study and selection of research methods, conducted data collection and data analyses, and was responsible for writing the manuscript. AC was involved in the selection of research methods, assisted with the interpretation of analyses, and provided feedback on multiple draft versions. NR was involved in the selection of research methods, processed raw data, assisted with the interpretation of analyses, and provided feedback on later draft versions. EA was involved in the conceptualization of the study and selection of research methods, assisted with the interpretation of analyses, and provided feedback on multiple draft versions. All authors thoroughly reviewed and approved the final manuscript before submission. EA is the guarantor.

During the preparation of this work the author(s) used Open AI’s ChatGPT Plus in order to edit the language with the aim to reduce word count and enhance the readability of the written manuscript.

### Ethical approval

This study involved human participants and was conducted in accordance with the Declaration of Helsinki. The study was approved by the Swedish Ethical Review Authority (approval number: 2024-04579-01). Written informed consent was obtained from all participants before data collection. Access to data is limited to the principal investigator (EA) and researchers involved in the analysis. In compliance with EU General Data Protection Regulation (GDPR) and the requirements of the Swedish Civil Contingencies Agency (MSB), data are stored on high-security data management platform at Lund University, Lund, Sweden.

### Patient consent for publication

Not applicable.

### Data availability statement

The data used in this study contains sensitive information about the study participants and they did not provide consent for public data sharing. The current approval by the Swedish Ethical Review Authority (approval number 2024-04579-01) does not include data sharing. A minimal data set could be shared by request from a qualified academic investigator for the sole purpose of replicating the present study, provided the data transfer is in agreement with EU legislation on the general data protection regulation and approval by the Swedish Ethical Review Authority. Contact information: Department of Health Sciences, Faculty of Medicine, Lund University Box 117, 221 00 Lund, Sweden; contact address; principal investigator: Eva Ageberg, Department of Health Sciences, Faculty of Medicine, Lund University, PO Box 117, SE-221 00 Lund, Sweden; Swedish Ethical Review Authority, Box 2110, 750 02 Uppsala, Sweden; phone: + 46 10 475 08 00.

## Acknowledgments

The authors would like to thank all participants for taking part in this study. Head acceleration and neck muscle strength data were collected at the Movement and Reality Lab (MoRe-Lab), Faculty of Medicine, Lund University. Finally, the authors thank physiotherapy students Rebecca Rose Christensson, Elsa Thomsen Olson, and Neo Spjut for their assistance with laboratory data collection.

