## Supplemental Data 1 for "Efficacy and acceptability of an end-user-informed neck resistance training program on outcomes related to sport-related concussion prevention in female football: A quasi-experimental pilot study"

### Supplemental material 2 – Acceptability Questionnaires

**Table 1.** Player acceptability questionnaire based on the Theoretical Framework of Acceptability (TFA).

| Construct | Definition | Statement/Questions |
|---|---|---|
| **Affective attitude** | "How an individual feels about the intervention." | 1. *I liked the exercise program.*<br>2. *I felt uncomfortable while engaging in the exercise program.* |
| **Burden** | "The amount of effort required to participate in the intervention." | 3. *This exercise program requires too much effort to perform.* |
| **Perceived effectiveness** | "The extent to which the intervention has achieved its intended purpose." | 4. *I could benefit from continuing to use this exercise program multiple times per week.* |
| **Intervention coherence** | "The extent to which the participant understands how the intervention works." | 5. *It is clear to me how this exercise program may affect outcomes related to prevention of sport–related concussion.* |
| **Self-efficacy** | "The participants' confidence that they can perform behavior(s) required to participate in the intervention." | 6. *I felt confident performing the exercise program.* |
| **Opportunity cost** | "The benefits, profits, or values that were given up to engage in the intervention." | 7. *Implementing this exercise program interfered with my other priorities.* |
| **Coping strategies** | "The process of managing demands (external or internal) that are appraised as taxing or exceeding the resources of the participant." | 8. *What issues might arise to prevent you from doing this exercise program?*<br>9. *How could issues mentioned above be overcome in order to continue doing this exercise program?* |

**Table 2.** Coach acceptability questionnaire based on the Theoretical Framework of Acceptability (TFA).

| Construct | Definition | Statement/Questions |
|---|---|---|
| **Affective attitude** | "How an individual feels about the intervention." | 1. *I liked the exercise program.*<br>2. *I felt uncomfortable while prescribing/supervising the exercise program.* |
| **Burden** | "The amount of effort required to participate in the intervention." | 3. *This exercise program requires too much effort to organize.* |
| **Perceived effectiveness** | "The extent to which the intervention has achieved its intended purpose." | 4. *Our team could benefit from continuing to use this exercise program multiple times per week.* |
| **Intervention coherence** | "The extent to which the participant understands how the intervention works." | 5. *It is clear to me how this exercise program may affect outcomes related to prevention of sport–related concussion.* |
| **Self-efficacy** | "The participants' confidence that they can perform behavior(s) required to participate in the intervention." | 6. *I felt confident prescribing/supervising the exercise program.* |
| **Opportunity cost** | "The benefits, profits, or values that were given up to engage in the intervention." | 7. *Implementing this exercise program may interfered with my other priorities.* |
| **Coping strategies** | "The process of managing demands (external or internal) that are appraised as taxing or exceeding the resources of the participant." | 8. *What issues might arise to prevent you from prescribing/supervising this exercise program?*<br>9. *How could issues mentioned above be overcome in order to continue doing this exercise program?* |
